# Lifespan EEG Reference Charts Reveal Aperiodic Confounds in Beta-Band Biomarkers Across Neurological and Neuropsychiatric Disorders

**DOI:** 10.64898/2026.09.19.26363458

**Authors:** Vardhan Paliwal, Alexander Moiseev, Sam M. Doesburg, Pengcheng Xi, Joel S. Winston, Mark P. Richardson, Roman Rodionov, Urs Ribary, Andrew Blaber, George Medvedev, Vasily A. Vakorin

## Abstract

Beta oscillations measured by scalp electroencephalography (EEG) are among the most prominent neural rhythms, implicated in motor control, cognition, and multiple neurological disorders, yet the absence of large-scale normative data and methodological inconsistency across studies has hindered the development of reliable reference frameworks necessary for clinical translation. These inconsistencies stem from two unresolved challenges: age-dependent neurophysiological changes that produce inherently non-linear beta trajectories across the lifespan, and the confounding influence of aperiodic 1/f activity in conventional beta analyses which changes independently with maturation and pathology. In this paper, we establish the first comprehensive lifespan reference charts for beta oscillations (*N* = 22,094, ages: 1-100 years) using complementary analytic approaches that isolate periodic oscillations from aperiodic background activity. We further quantify disorder-specific deviations in beta power and frequency across seven neurological and neuropsychiatric disorders. Beta power showed a non-linear, tri-phasic trajectory, increasing through childhood and adolescence, peaking around age ~50 years, and declining in later life. Unadjusted and aperiodic-adjusted beta frequency showed opposing developmental trajectories; adjusted frequency revealed a previously uncharacterized adolescent dip, reaching minimum around ages 12–13 years, consistent across all brain lobes and both sexes. Beta power was reduced across most clinical groups, with effect sizes varying by disorder and brain lobe; schizophrenia spectrum disorders were a notable exception, showing reduced absolute but increased aperiodic-adjusted beta power — a distinction undetectable without aperiodic decomposition. These reference charts demonstrate that aperiodic 1/f activity does not merely scale beta measurements but reverses their apparent developmental trajectory — a systematic bias that, left unaccounted for, fundamentally misrepresents how beta frequency matures across the lifespan and obscures disorder-specific oscillatory signatures in clinical populations.

## 1. Introduction

Beta oscillations (13–30 Hz) are among the most prominent and functionally significant neural rhythms in the human brain. During active behavior, these oscillations show characteristic event-related dynamics: power decreases (desynchronization) during movement preparation and execution, followed by post-movement rebound during motor termination and steady-state maintenance [1, 2]. This task-dependent modulation supports the “status quo” hypothesis, whereby beta oscillations promote maintenance of current sensorimotor and cognitive states while suppressing competing actions [3], with further roles in top-down attentional control, working memory, and cognitive control [4–6]. Beyond the task-evoked dynamics, spontaneous resting-state beta, the dominant expression of beta-band activity in routine clinical EEG, reflects the baseline inhibitory tone of cortical networks. Resting beta power directly indexes cortical GABAergic inhibitory tone [7–9], exhibits high intra-individual reliability (ICC > 0.90) [10], and independently predicts motor function across the lifespan [11, 12]. Critically, resting beta undergoes systematic changes across development and aging [13, 14], and shows disorder-specific alterations across a broad range of neurological and neuropsychiatric conditions [15, 16].

Despite their functional significance, beta oscillations remain one of the least understood neural rhythms, particularly regarding their developmental trajectories across the lifespan. The contradictions in the existing literature cluster around three sources of inconsistency. First, studies disagree on the direction and shape of the developmental trajectory: some studies report beta power increases during adolescence and young adulthood [14], while others report stable levels across childhood [17], non-linear patterns with increases in middle age [18], or decreases during childhood and adolescence [19]. Second, regional specificity complicates interpretation: sensorimotor beta power increases with healthy aging and predicts motor function decline [11], yet task-related beta modulation decreases in older adults despite elevated resting beta [20, 21]. Third, conflation of beta sub-bands obscures developmental patterns: low beta (12–20 Hz) and high beta (20–30 Hz) follow entirely different trajectories, with high beta peaks present in 99.5% of infants under 12 months while low beta emerges gradually, appearing in only 48% of infants by 18–20 months [22]. Beta frequency development remains similarly unresolved, with scattered reports of increases during childhood [23] or no developmental correlation at all [24].

These contradictions likely reflect the inherent complexity of beta-band activity generation and maturation. Beta oscillations emerge from distributed cortical-subcortical networks involving motor cortex, basal ganglia, and thalamocortical circuits [3, 25]. Multiple developmental processes including myelination, inhibitory circuit maturation, and thalamocortical refinement unfold at different rates across brain regions and continue into adulthood [26], creating inherently non-linear developmental trajectories. These processes unfold non-uniformly across development, creating trajectories that are highly individualized even among healthy individuals.

Biological complexity alone, however, cannot fully account for the contradictory findings in the literature. A fundamental methodological limitation further confounds the interpretation of beta-band activity development: conventional spectral analysis conflates periodic beta oscillations with aperiodic 1/f activity [27]. This aperiodic component reflects non-oscillatory neural processes arising from asynchronous neuronal firing and is a proxy to the balance between excitation and inhibition within neural circuits [28]. Functionally, 1/f activity is modulated during cognitive tasks [29], altered by pharmacological interventions [8, 30], and varies with arousal states [31]. It also undergoes systematic changes across development and in neurological disorders [32, 33]. The aperiodic exponent and offset decrease through childhood and adolescence, representing progressive power spectrum flattening. Conventional bandpass filtering cannot separate these aperiodic changes from periodic beta oscillations, potentially misattributing broadband changes to specific oscillatory alterations [32].

The combination of inherent developmental complexity and methodological confounds has direct consequences for clinical interpretation. Beta-band alterations have been reported across multiple neurological disorders, but findings remain inconsistent and difficult to interpret without normative baselines. Disorders associated with neurodegeneration show opposing beta profiles depending on disease stage and circuit involvement. In Parkinson’s disease, early-stage pathology is associated with elevated resting sensorimotor cortical beta power [34], while later-stage patients show widespread cortical beta power reductions alongside general spectral slowing [15, 35]. Dopaminergic medication partially normalizes these cortical reductions [15], though subcortical beta dynamics show opposing medication effects. In contrast, Alzheimer’s disease shows consistent cortical beta power reductions that predict cognitive decline [36], reflecting a predominantly hypo-beta profile from early disease stages. Critically, healthy aging itself produces elevated sensorimotor beta power that overlaps with early Parkinsonian features [37], meaning that both beta increases and decreases fall within the range of normal age-related variation. Without a normative baseline spanning the full human lifespan, it remains difficult to determine whether observed beta changes in an individual patient reflect pathology or age-appropriate variation, fundamentally limiting the clinical diagnostic utility of beta-band activity.

Here we address these limitations using a large-scale clinical EEG dataset (*N*=22,094; ages 1–100 years). We establish normative lifespan trajectories for beta power and frequency using both conventional bandpass filtering and spectral parameterization, directly quantifying how aperiodic 1/f activity confounds traditional beta measurements across development. We further characterize sex-stratified trajectories across six cortical lobes, and extend beyond case-control comparisons by quantifying disorder-specific deviations from age-appropriate normative expectations across seven neurological and neuropsychiatric conditions, enabling identification of disorder-specific signatures while controlling for normal developmental and aging effects.

## Methods

### Patient Demographics

We analyzed a large dataset of routine clinical electroencephalography (EEG) recordings collected between 2010 and 2018 across four public hospitals within the Fraser Health Authority, British Columbia, Canada. No new human subjects were recruited or tested. The original data were collected under ethical approval by Simon Fraser University and Fraser Health Authority on 1^*st*^ April 2022 (protocol number: H18-02728). All data were anonymized/de-identified prior to analysis. Patient demographic information, including age and sex, was incorporated into the analyses, and EEG recordings were selected without bias. Males and females were analyzed separately, and patients were classified as inpatients or outpatients according to the criteria of the Canadian Institute for Health Information (CIHI). The cohort encompassed a culturally and socioeconomically diverse population.

Our data consists of two patient population: outpatients and inpatients. Outpatients were defined as individuals who visited the hospital for clinical assessments but were not admitted. This group comprised *N* = 22,094 patients, including 10,939 males (mean age: 43.3 ± 23.1 years) and 11,154 females (mean age: 44.1 ± 22.2 years). Inpatients were defined as individuals who spent at least one night in the hospital and received acute care, excluding those seen exclusively in emergency departments. Inpatient cases were categorized into Case Mix Groups (CMGs) defined by the Canadian Institute for Health Information (CIHI) using the International Statistical Classification of Diseases and Related Health Problems, 10th Revision, Canada (ICD-10-CA). Within this categorization, we focused on the diagnostic category of mental disorders, and identified seven clinical groups based on the highest sample size, with the threshold of at least 50 male or female inpatients. Demographic characteristics of the inpatient groups are provided in Table 1. Inpatients were further stratified by comorbidity scores (0–4), reflecting overall health status, with 0 indicating good health and 4 denoting severe comorbidity burden.

**Table 1.** Demographics of Inpatients’ Cohort.

| Mental Disorder | Sex | Age Range (years) | Number of Inpatients | Mean Age ± SD (years) |
| --- | --- | --- | --- | --- |
| Seizure Disorder | Male | 1 - 93 | 550 | 49.4 ± 26.2 |
| | Female | 1 - 97 | 454 | 53.5 ± 26.5 |
| Status Epilepticus | Male | 1 - 94 | 157 | 52.2 ± 21.3 |
| | Female | 1 - 90 | 152 | 52.4 ± 16.9 |
| Organic Mental Disorder | Male | 20 - 95 | 186 | 71.8 ± 14.2 |
| | Female | 33 - 104 | 136 | 76.7 ± 13.4 |
| Dementia | Male | 40 - 95 | 88 | 75.2 ± 10.9 |
| | Female | 43 - 97 | 59 | 77.3 ± 11.3 |
| Substance Abuse | Male | 20 - 76 | 68 | 49.8 ± 14.3 |
| | Female | 16 - 77 | 27 | 47.4 ± 16.7 |
| Schizophrenia | Male | 17 - 73 | 76 | 37.3 ± 15.9 |
| | Female | 16 - 80 | 55 | 38.2 ± 17.7 |
| Schizotypal Disorder | Male | 11 - 82 | 103 | 35.0 ± 17.2 |
| | Female | 14 - 84 | 108 | 42.3 ± 18.1 |

### EEG Preprocessing

EEG recordings were acquired using standardized hardware and firmware across all sites. Each recording station was equipped with a Natus Xltek EEG32U amplifier and gold-cup electrodes. Data were collected using the international 10–20 system from 20 channels: FP1, FPZ, FP2, F3, F4, F7, F8, FZ, T3, T4, T5, T6, C3, C4, CZ, P3, P4, PZ, O1, and O2. The original sampling frequency was maintained at either 500 or 512 Hz consistently within each recording.

Preprocessing followed a standardized pipeline. First, raw EEGs were recordings converted from the Natus proprietary format to European Data Format (EDF) and then anonymized using the PyEDFlib library [38] in Python. A zero-phase, overlap-add finite impulse response band-pass filter (0.5–55 Hz with Hamming window) was applied using MNE-Python [39] to retain canonical brain rhythms while excluding noise above the 60 Hz powerline frequency. All recordings were resampled to 256 Hz for standardization. Non-neural segments, including flat intervals (digital zeros with a minimum peak-to-peak threshold of 1e-6), photic stimulation, and hyperventilation procedures, were removed. The resulting dataset comprised 6-minute, 20-channel, sensor-space EEG recordings.

### Source Reconstruction and EEG Spectral Power Estimation

We reconstructed EEG source activity at 148 cortical regions of interest (ROIs; 74 per hemisphere) [40] defined by the Destrieux atlas [41]. Source reconstruction employed a standardized fsaverage template MRI from FreeSurfer with standard electrode positions for the 10-20 montage, consistent with established practice for large-scale retrospective clinical EEG cohorts where individual MRIs are unavailable. This template was applied uniformly across all age groups. Each ROI was further grouped into six cortical lobes: insular, limbic, frontal, occipital, parietal, and temporal. Source reconstruction was carried out using a beamforming approach [42]. Forward solutions (lead fields) [43] were computed for each cortical source using the MNE-Python library, based on a three-layer boundary element model (BEM) of the head. Standard conductivity values were assigned to the inner skull (0.3 S/m), outer skull (0.006 S/m), and scalp (0.3 S/m). Neural time series were then reconstructed using a scalar single-source minimum variance beamformer [43]. Beamformer weights were derived to minimize output variance while enforcing unit gain at the target source, thereby attenuating contributions from other cortical regions and extraneous noise. To mitigate depth-related biases intrinsic to beamformer reconstructions, the resulting source time series were normalized by the root mean square (RMS) of the projected sensor noise covariance. This procedure produces dimensionless pseudo-Z scores that index the local signal-to-noise ratio, facilitating unbiased comparisons across cortical regions irrespective of source depth.

Power spectral density was estimated for each source time series using Welch’s method [44], as implemented in the SciPy library [45]. Spectra were computed over the 1–55 Hz range with a frequency resolution of 0.5 Hz, resulting in 109 frequency bins. The resulting dataset consisted of spectral power estimates for 148 cortical sources across these 109 frequency points.

From the source-reconstructed EEG spectral power, beta oscillatory parameters (frequency and power) were extracted using two complementary approaches: band-pass filtering and spectral parameterization into periodic and aperiodic components. In the band-pass filtering approach, beta-band activity was defined within the 13–30 Hz range. A band-pass filter was applied to isolate this range, and the frequency with maximal power was identified as the Unadjusted Beta Frequency. The corresponding power value was defined as Absolute Beta Power, expressed in (pseudo-Z)^2^/Hz. This method captured total beta-band activity within the EEG spectrum. In the spectral parameterization approach, we employed the toolbox described in [27], which decomposes spectral power into an aperiodic (1/f) component and Gaussian distributions modeling periodic oscillatory peaks. Beta oscillations were defined within the 13–30 Hz range; if multiple peaks were present, the one with the highest power was identified as the Adjusted Beta Frequency, with the corresponding power defined as Adjusted Beta Power (arbitrary units, a.u.). This approach isolated periodic beta oscillations while accounting for the aperiodic background. For both methods, beta frequency and power were computed for each ROI and subsequently averaged across ROIs within each cortical lobe to obtain representative lobe-wise values.

### Workflow of Analyses

Using outpatient EEGs, we generated scatterplots of aperiodic adjusted and unadjusted beta oscillatory parameters, stratified across six cortical lobes. To establish developmental trajectories across the lifespan, we applied locally weighted scatterplot smoothing (LOWESS) from the Statsmodels library [46], a non-parametric regression technique that produces robust local polynomial fits while minimizing the influence of outliers.

We also assessed the differences in the normative developmental trajectories across brain lobes and sex. To assess regional heterogeneity and sex differences in lifespan trajectories, we computed pairwise differences between all six cortical lobes for each spectral parameter within individual participants. This produced distributions of inter-regional differences (e.g., occipital minus temporal values) across the cohort. Effect sizes (Cohen’s *d*) were then calculated for these distributions relative to zero difference. Results were summarized in a 6×6 heatmap for each parameter: the lower triangle cells represented effect sizes for inter-regional differences in male trajectories, the upper triangle cells represented those for females, and the diagonal cells captured sex differences within each lobe. This provided an overall map of regional variability, quantifying both inter-regional and sex-related differences while accounting for individual-specific scaling factors.

Normative trajectories were constructed from outpatient EEGs on the basis of two complementary advantages. First, this sample provides continuous lifespan coverage spanning infancy to advanced age — a demographic range that purpose-recruited healthy control cohorts cannot feasibly achieve. Second, the large sample size affords statistical robustness: although the population is clinically heterogeneous, encompassing individuals with normal findings, non-specific abnormalities, and defined neurological conditions - individual pathological contributions are effectively attenuated when estimating population-level age trajectories, yielding stable reference standards representative of the broader clinical population encountered in routine EEG practice. Reference standards derived in this manner carry direct translational relevance, providing a valid benchmark against which deviations in clinical groups can be meaningfully quantified.

Next we aimed to quantify alterations in oscillatory parameters among clinical groups. To achieve this, inpatient beta parameters were projected onto outpatient reference charts and the differences were modeled as individual deviations from the outpatient trajectories. The distribution of these deviations was examined across cortical lobes and diagnostic categories. Statistical significance was assessed using one-sample t-tests (tested against a mean of zero), and effect sizes were calculated with Cohen’s *d*. Further, we quantified the associations between individual deviations in beta frequency/power and comorbidity scores. To quantify these, we used OrderedModel function in Statsmodels, which implements ordinal logistic regression. This model estimated odds ratios, reflecting the likelihood of higher comorbidity scores as a function of deviations in beta parameters, and provided corresponding p-values.

Effect size (Cohen’s *d*) was the primary metric of clinical deviation magnitude, as it quantifies the practical significance of group differences independently of sample size. Consistent with ASA guidelines on statistical inference [47, 48], p-values were treated as continuous descriptive statistics and interpreted in conjunction with effect sizes rather than as binary arbiters of significance: a p-value threshold does not demarcate findings that are real from those that are not, and should not be used as such. Following Benjamini-Hochberg false discovery rate correction across brain lobes and spectral parameters, *p* < 0.1 was used as a secondary screening heuristic to organize reported findings; it governed what was reported, not what was concluded. The magnitude and direction of Cohen’s *d* remains the basis for all clinical interpretations. The workflow is presented in Figure 1.

**Figure 1:**
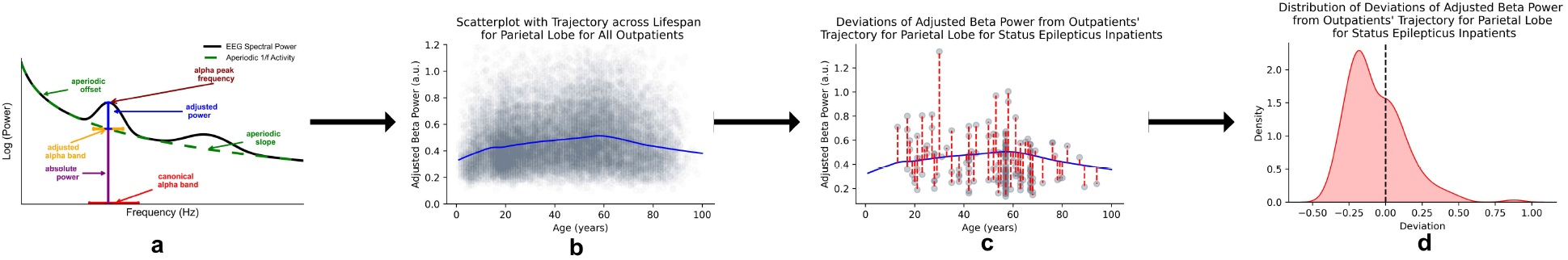
Workflow of analysis for adjusted beta power for parietal lobe. **b** The trajectory across lifespan is generated based on scatterplot distribution of outpatients’ adjusted beta power. Adjusted beta power of status epilepticus inpatients’ parietal lobe were projected onto this trajectory. **c** This population had higher proportion of their beta power below the outpatient reference trajectory. **d** Distribution of deviations from the outpatient reference chart indicating a reduced adjusted beta power in parietal lobe for status epilepticus inpatients.

### Results

### Beta Power Increases Non-Linearly Until Mid-Adulthood Followed by Decline in Later Years

Lifespan reference charts for both absolute and 1/f-adjusted beta power revealed a consistent non-linear trajectory across sex and brain lobes. Figure 2 shows the normative reference trajectories for beta power, stratified by estimation method, sex, and brain lobe. Both absolute and 1/f adjusted power increased non-linearly through childhood and adolescence, peaked around age ~ 50, and declined in later years. This pattern was consistent across sex and brain lobes. We also observed that female absolute and 1/f adjusted beta power were higher than male values across the lifespan (Fig. 2**b,e**), a difference formally confirmed by medium effect sizes (Cohen’s *d* < 0.5) in cross-sex trajectory comparisons (Fig. 3**a,b**). Moreover, both beta power trajectories exhibited lobe-dependent differences, with effect sizes varying from low-to-high across brain lobes (Fig. 3**a,b**).

**Figure 2:**
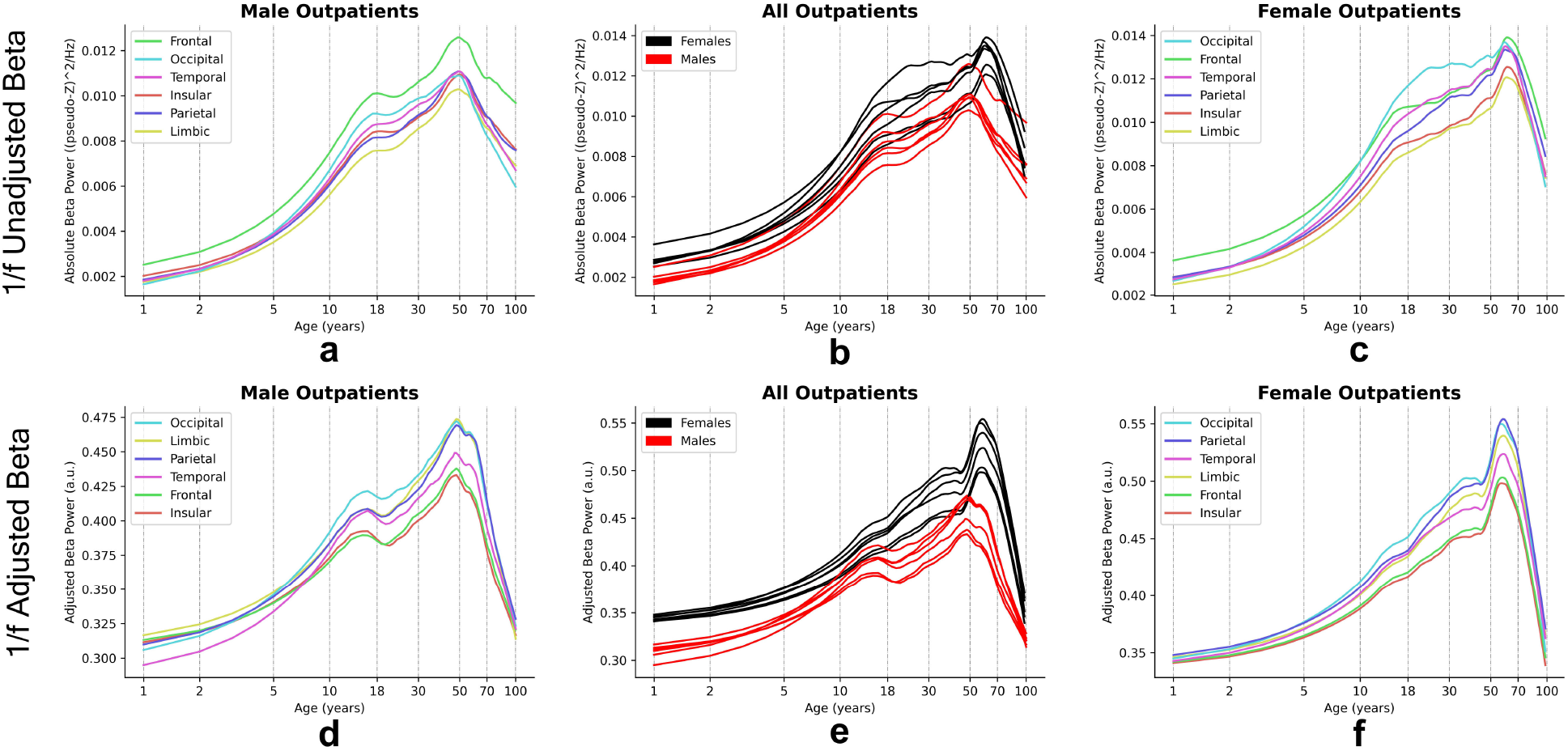
Changes in beta power as a function of age across lifespan due to variation in three factors: brain lobes, sex, and estimation methods. The lifespan trajectories are computed using scatterplot averaging. The age on the x-axis is represented using natural logarithmic scale. Both absolute and adjusted beta power showed similar lifespan trajectories: characterized by a non-linear increase through childhood and adolescence until mid adulthood years, where beta power reached its peak (age ~ 50 years), and then declined in later years. **(a)** Lifespan reference charts for absolute beta power for males across six brain lobes. **(b)** Lifespan reference charts for absolute beta power for males and females compared across six brain lobes. **(c)** Lifespan reference charts for absolute beta power for females across six brain lobes. **(d)** Lifespan reference charts for adjusted beta power for males across six brain lobes. **(e)** Lifespan reference charts for adjusted beta power for males and females compared across six brain lobes. **(f)** Lifespan reference charts for adjusted beta power for females across six brain lobes.

**Figure 3:**
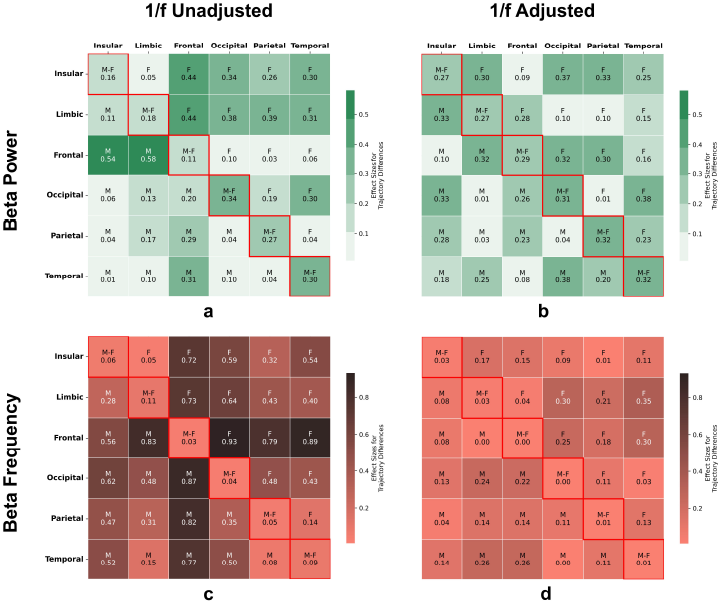
Comparison of lifespan reference trajectories across sex and brain lobes for unadjusted and 1/f-adjusted beta parameters. Each heatmap summarizes pairwise effect sizes (Cohen’s *d*) between regional and sex-stratified lifespan trajectories for one beta parameter. The lower triangle represents inter-regional comparisons for males, the upper triangle for females, and the diagonal cells (red borders) reflect sex differences within corresponding brain lobes. Each cell displays the effect size value and comparison type (M: between male trajectories; F: between female trajectories; M-F: between sexes). Darker colors denote larger effect sizes. Absolute and adjusted beta powers are displayed on matched color scales, as are unadjusted and adjusted beta frequencies, to facilitate direct visual comparison between estimation approaches. **a** Absolute beta power: inter-regional differences showed low-to-medium effect sizes in both sexes, while sex differences within lobes were low. **b** Adjusted beta power: inter-regional differences were consistently low, whereas sex differences reached medium effect sizes. **c** Unadjusted beta frequency: inter-regional differences were medium-to-high, reflecting substantial regional heterogeneity, while sex differences remained very low. **d** Adjusted beta frequency: both inter-regional and sex differences showed low effect sizes, indicating greater homogeneity across regions and sexes following aperiodic adjustment.

Both absolute and adjusted beta power appeared to peak at different ages for males and females: around 50 years in males versus 65 years in females. Further, despite similar developmental patterns observed in absolute and adjusted beta power trajectories, a divergence emerged in late-life trajectories between the two estimates. Adjusted beta power displayed a pronounced decline in older age, with values in centenarians approaching those observed in early childhood, creating an almost ‘circular’ lifespan pattern (Fig. 2**d-f**). In contrast, absolute beta power, while declined, maintained distinct adult-like characteristics throughout late life and did not regress to infantile levels (Fig. 2**a-c**).

The consistency of this trajectory across methods, sex, and brain lobes establishes a robust normative reference for beta power against which disorder-related deviations can be assessed.

### Beta Frequency Shows Divergent Lifespan Trajectories After 1/f Adjustment

Lifespan reference charts for beta frequency revealed strikingly different developmental trajectories depending on whether aperiodic activity was accounted for. Figure 4 shows the normative reference trajectories for beta frequency, stratified by estimation method, sex, and brain lobes. Unadjusted beta frequency remained constant during childhood, followed by an increase in adolescence, plateaued until the mid-adulthood years (age ~ 50 years), and then decreased for the remaining lifespan (Fig. **4a-c**). This pattern was consistent across the brain lobes and sex. Visually, we observed high inter-regional variability in unadjusted beta frequency for both male and female trajectories, which was confirmed with high effect sizes (Cohen’s *d* > 0.8) (Fig. 3**c**).

**Figure 4:**
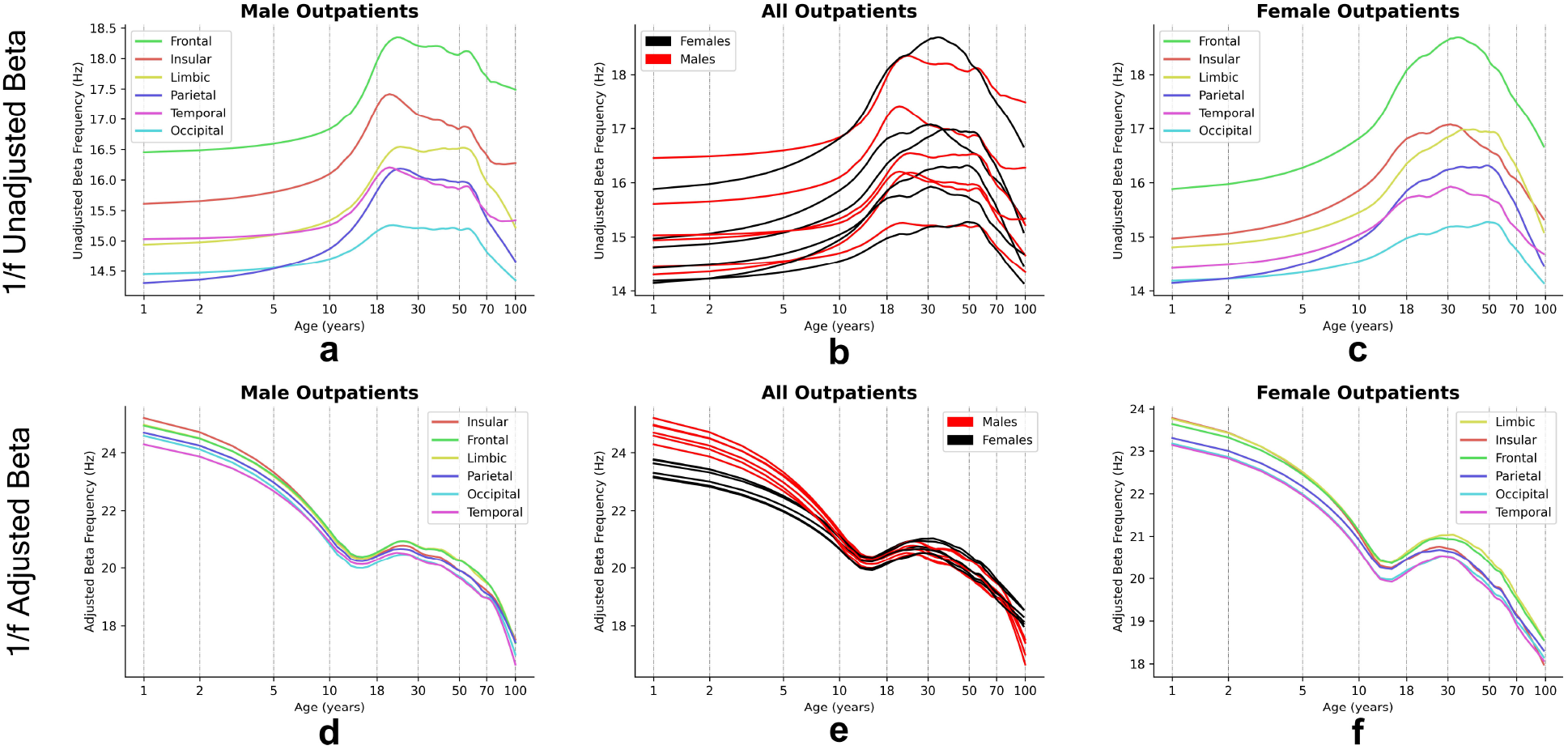
Changes in beta frequency as a function of age across lifespan due to variation in three factors: brain lobe, sex, and estimation method. The lifespan trajectories are computed using scatterplot averaging. The age on the x-axis is represented using natural logarithmic scale. Unadjusted beta frequency remained constant in childhood, followed by an increase in adolescence years, plateaued until mid-adulthood years, and then declined in the later years of the lifespan. **(a)** Lifespan reference charts for unadjusted beta frequency for males across six brain lobes. **(b)** Lifespan reference charts for unadjusted beta frequency for males and females compared across six brain lobes. **(c)** Lifespan reference charts for unadjusted beta frequency for females across six brain lobes. Adjusted beta frequency decreased in childhood, followed by a brief recovery period of increase in adolescence years until young adulthood (age ~ 30 years), and then again decreased throughout the remaining lifespan. **(d)** Lifespan reference charts for adjusted beta frequency for males across six brain lobes. **(e)** Lifespan reference charts for adjusted beta frequency for males and females compared across six brain lobes. **(f)** Lifespan reference charts for adjusted beta frequency for females across six brain lobes.

Adjusted beta frequency in contrast showed a directionally opposite developmental trajectory to unadjusted frequency. Adjusted beta frequency decreased in childhood, followed by a brief recovery period of increase throughout the adolescence until ~30 years, and then progressively decreased through the later years of life. This contrasting pattern was consistent across both brain lobes and sex. Visually, the trajectories appear to be similar both across sex and across brain regions, and this was further confirmed with low effect sizes (Cohen’s *d* < 0.2) for both inter-region and inter-sex differences (Fig. 3**d**). Unadjusted and aperiodic-adjusted beta frequency thus traced opposing developmental trajectories across the lifespan.

### Characterization of the Dip in the Adjusted Beta Frequency Trajectory

A striking and unexpected finding from our normative beta frequency charts was the trajectory pattern of adjusted beta frequency. It decreased throughout childhood, increased during adolescence into young adulthood (age ~30 years), then decreased again across the remaining lifespan, creating a distinctive dip during late childhood through early adulthood. This dip was visually consistent across brain lobes and sex. However, it remained unclear whether this reflected a genuine developmental feature or an artifact of the LOWESS smoothing procedure, which can produce spurious local minima in age ranges with sparser sampling or greater individual variability — both characteristics of the adolescent period in clinical EEG datasets. Critically, a methodological artifact would be expected to vary arbitrarily across brain regions and between sexes, whereas a genuine developmental feature should manifest consistently, with reproducible timing, depth, and duration.

Given that this dip was unexpected and not part of the original analysis plan, we conducted a series of post hoc analyses to verify whether it reflected a genuine developmental feature or a smoothing artifact. We first smoothed lifespan trajectories using LOWESS with log-transformed age values. A genuine adolescent dip characterized by an initial decrease, a local minimum, and subsequent recovery requires at least two critical points, which a cubic polynomial can capture but a quadratic or linear model cannot. We therefore fitted linear, quadratic, and cubic polynomial models to log-transformed age data, reasoning that a cubic model would provide a significantly better fit than simpler alternatives. Models were compared using Akaike Information Criterion (*AIC*) (Δ*AIC* > 2 indicating significant improvement). When cubic models provided superior fits, we detected local minima characterizing the dip, using Scipy’s argrelextrema function [45], prioritizing minima occurring within the adolescent age range (8-25 years). For each detected adolescent dip, we quantified the following dip characteristics: (1) dip depth (baseline minus minimum frequency in Hz), (2) dip duration (dip onset to recovery in years), and (3) dip timing (age at minimum in years). Baseline frequencies were estimated using averaged frequency values from childhood (1-8 years) and younger-adulthood (25-35 years) periods, with recovery points identified through derivative analysis of the trajectories. Further, bootstrap resampling (n=1000) was employed to generate 95% confidence intervals for dip depth and timing estimates. Finally, we applied cross-region consistency check for characterizing the dip characteristics in terms of mean and standard deviations for dip depth and dip timing, separately for males and females. This approach enabled the quantification of the adolescent dip across multiple brain regions while accounting for inter-individual variability in developmental trajectories.

Cubic models consistently outperformed linear and quadratic models across all brain regions for both sexes (Δ*AIC* > 10; Table 2), confirming the existence of the adolescent dip. We then characterized three key dip properties: depth, timing, and duration. Dip depth was approximately 2 Hz below baseline frequency across all brain lobes in both sexes (Table 2). Dip timing, the age at minimum frequency, occurred at 13.0 years in males and 12.0 years in females, consistent across all brain lobes (Table 2). Dip duration was stable at approximately 20 years in females across brain lobes, but showed greater variability in males. Cross-region analysis confirmed that all six brain lobes in both sexes exhibited Δ*AIC* > 10 for cubic versus quadratic models, demonstrating the global consistency of this developmental feature in adjusted beta frequency normative trajectories (Table 3).

**Table 2.** Adolescent Dip Characteristics in Adjusted Beta Frequency Trajectories across Brain Lobes and Sex.

| Brain Region | Sex | $\Delta AIC$<br>(quad→cubic) | Dip Timing<br>(years) | 95 % CI for<br>Dip Timing<br>(years) | Dip Depth<br>(Hz) | 95 % CI for<br>Dip Depth<br>(Hz) | Baseline<br>Frequency<br>(Hz) | Recovery<br>Age<br>(years) | Dip<br>Duration<br>(years) |
| --- | --- | --- | --- | --- | --- | --- | --- | --- | --- |
| Insular | Male | 63.2 | 13.0 | [12.0, 14.0] | 2.11 | [1.72, 2.37] | 22.27 | 24.0 | 17 |
|  | Female | 179.8 | 12.0 | [12.0, 13.0] | 2.17 | [1.81, 2.49] | 22.11 | 26.0 | 19 |
| Frontal | Male | 83.6 | 13.0 | [12.0, 14.0] | 1.88 | [1.66, 2.23] | 22.12 | 38.0 | 31 |
|  | Female | 231 | 12.0 | [12.0, 13.0] | 2.03 | [1.74, 2.32] | 22.16 | 26.0 | 19 |
| Limbic | Male | 82.5 | 13.0 | [12.0, 14.0] | 2.01 | [1.80, 2.36] | 22.15 | 38.0 | 31 |
|  | Female | 247.6 | 12.0 | [11.0, 13.0] | 2.14 | [1.86, 2.45] | 22.24 | 27.0 | 20 |
| Occipital | Male | 78.3 | 13.0 | [12.0, 14.0] | 2.07 | [1.68, 2.27] | 21.84 | 24.0 | 17 |
|  | Female | 224.4 | 12.0 | [12.0, 13.0] | 2.15 | [1.82, 2.45] | 21.83 | 28.0 | 21 |
| Parietal | Male | 73.6 | 13.0 | [12.0, 14.0] | 1.84 | [1.63, 2.24] | 21.85 | 38.0 | 31 |
|  | Female | 172.1 | 12.0 | [12.0, 13.0] | 1.93 | [1.67, 2.23] | 21.87 | 26.0 | 19 |
| Temporal | Male | 69.4 | 13.0 | [12.0, 15.0] | 1.64 | [1.40, 2.05] | 21.57 | 38.0 | 31 |
|  | Female | 212.6 | 12.0 | [12.0, 13.0] | 2.08 | [1.76, 2.39] | 21.77 | 28.0 | 21 |

- $\Delta AIC$ values represent improvement of cubic over quadratic models (if $\Delta AIC > 10$ , indicator of strong evidence for cubic fit) - Dip duration calculated as recovery age minus onset age - Confidence intervals derived from bootstrap resampling ( $n = 1000$ ) - Recovery age defined as return to within 95% of baseline frequency

**Table 3.** Summary for Adolescent Dip Characteristics for Adjusted Beta Frequency.

| Sex | Mean $\Delta AIC$<br>(quad→cubic) | Mean Dip Depth<br>(Hz) | Mean Dip Duration<br>(years) | Mean Recovery Age<br>(years) | Mean Dip Timing<br>(years) |
| --- | --- | --- | --- | --- | --- |
| Males | 75.1 ± 7.5 | 1.93 ± 0.16 | 26.3 ± 7.5 | 31.7 ± 7.5 | 13.0 ± 0.0 |
| Females | 211.3 ± 31.4 | 2.09 ± 0.08 | 19.8 ± 1.0 | 26.8 ± 1.0 | 12.0 ± 0.0 |

These results establish the adolescent dip in adjusted beta frequency as an intrinsic feature of neurodevelopment rather than a methodological artifact.

### Beta Oscillations Show Disorder- and Region-Specific Alterations

Building on our normative lifespan reference charts, we next investigated alterations in beta power and frequency across multiple neurological and psychiatric disorders. Figure 6 shows the characterization of beta power alterations across neurological and neuropsychiatric disorders, stratified by sex and brain lobes. Absolute beta power showed a robust reduction in clinical groups compared to our reference trajectories (Fig. 6**a,c(i)**). This reduction was consistent across diagnostic categories, sex, and brain lobes, and appeared with low-to-high effect size with seizure disorder, status epilepticus, and organic mental disorders showing the largest effects (Fig. 6**a,c(ii)**). Adjusted beta power also showed reductions in most disorders, with the exception of schizophrenia spectrum disorders, where an increase in adjusted beta power was observed. The effects of reduction in adjusted beta power were strongest in organic mental disorder (Cohen’s *d* > 0.4). Together, these findings suggest that beta power reduction is a common feature of neurological illness, but its severity is shaped by diagnostic category and brain lobe.

**Figure 5:**
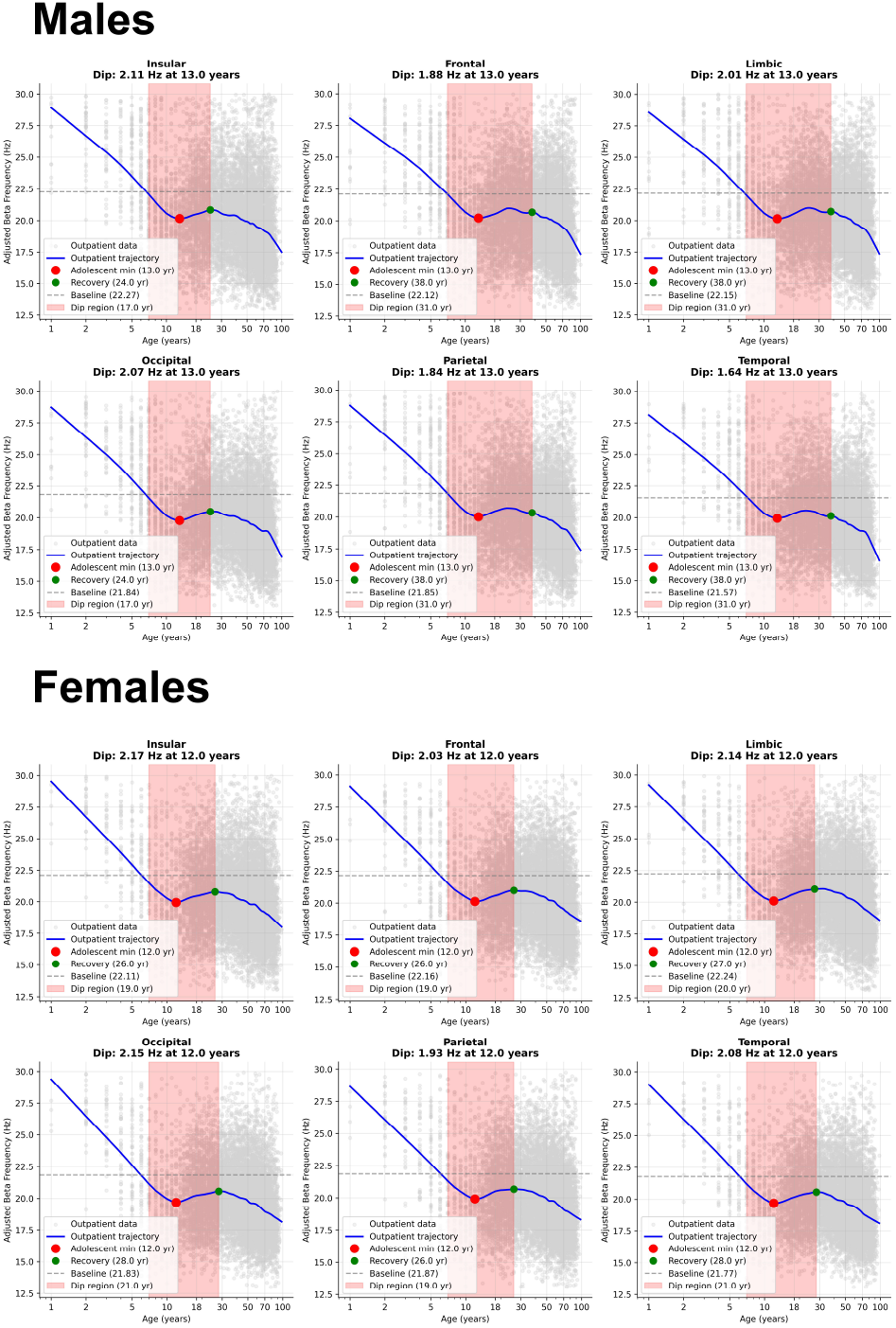
Quantification of the Adolescent Dip in Adjusted Beta Frequency. Dip characteristics are quantified separately for each brain lobe and sex. Each subplot represents age on the x-axis on a natural log scale to better represent the developmental changes in the early stages of lifespan. Y-axis represents adjusted beta frequency in Hz. Cubic model best explained the outpatients adjusted beta frequency data, confirming that the adolescent dip was a feature of our data. Next, scatterplot trajectories were generated based on LOWESS smoothing. The dip regions were identified and visualized with the following markers: adolescent minimum (age at dip minimum), recovery age (age where adjusted beta frequency returned to the baseline value), and dip region (duration of this dip). Each subplot confirms that the location of the dip is around the adolescent period, with dip minimum occurring around 12-13 years. The duration of the dip varied across brain lobes and between sex. Overall, the existence of the dip was a global effect present across all brain-regions with some variability in the dip parameters across brain lobes and across sex.

**Figure 6:**
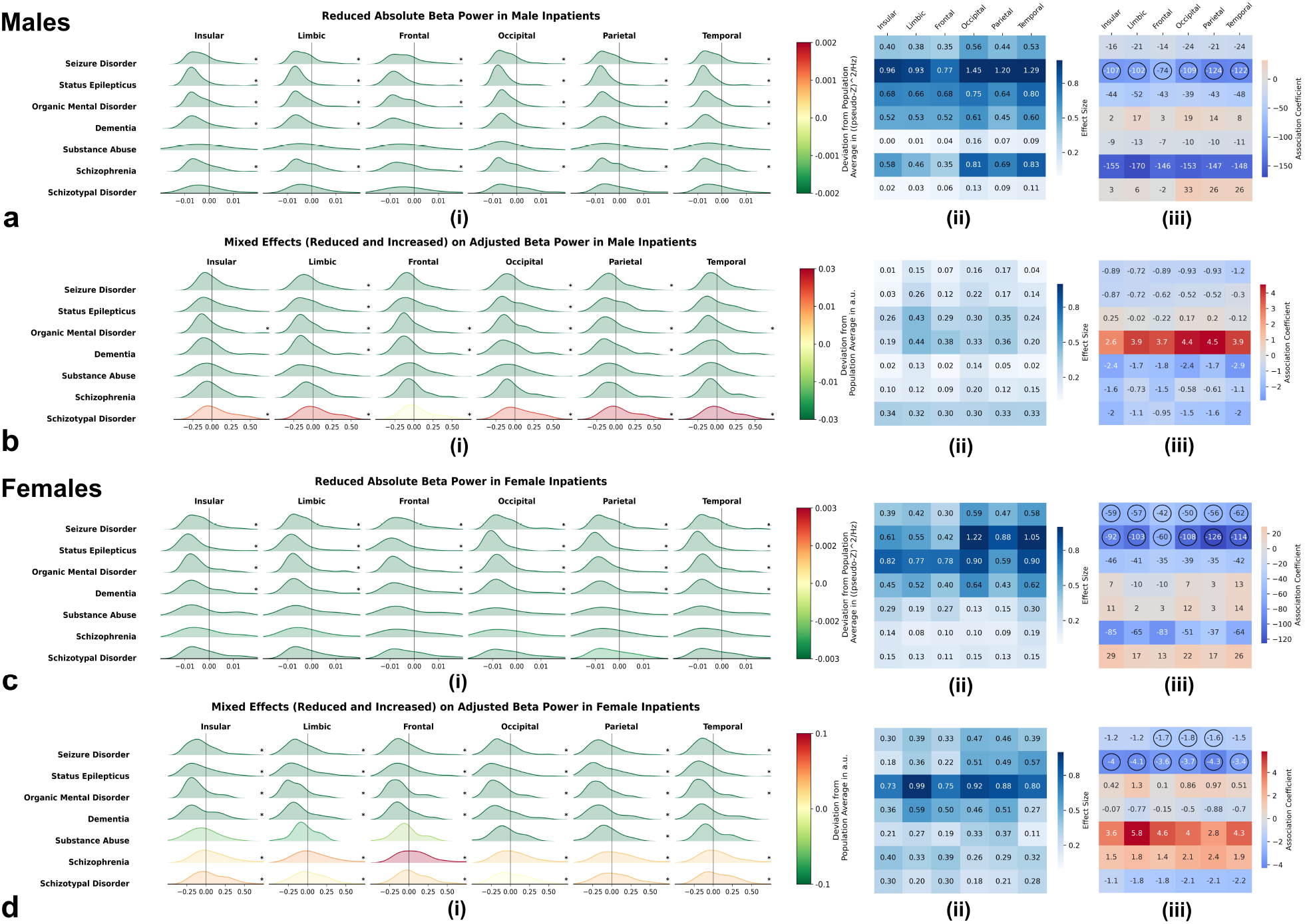
Alterations in Beta Power across Clinical Groups are Disorder-Specific. Panels a-d represent the characterization of beta power for clinical groups representing neurological disorders, stratified by sex, brain regions, and estimation method. **a-d(i)** Distributions of deviations in beta power from lifespan reference charts. Green distributions represent negative deviations indicating reduced beta power while red distributions represent positive deviations indicating increased beta power in neurological disorders, with statistical significance marked by (*). Reductions in both absolute and adjusted beta powers were observed in most clinical groups (*p* < 0.1), except schizophrenia spectrum, which showed increased adjusted beta power. These effects were consistent across sex and brain lobes. **a-d(ii)** Demonstrate the effect sizes of these deviations, with darker colors indicating stronger effects. The magnitude of these effects manifested with low-to-high effect sizes, varying across clinical groups. Seizure disorder and status epilepticus showed the largest effect, while schizophrenia spectrum disorders showed smallest, still significant reductions. **a-d(iii)** Display correlations between deviations in beta power and comorbidity scores with statistically significant correlations marked by circles. Negative correlations are shown as blue cells, while positive correlations are displayed in red cells. Significant negative correlations were observed, particularly in seizure disorder and status epilepticus inpatients (*p* < 0.1). The negative correlations implied that inpatients with these disorders showed smaller reductions in both absolute and adjusted beta powers with increasing comorbidity scores.

We also examined whether comorbidity scores correlated with these deviation scores. We observed that higher comorbidity was associated with smaller negative deviations in beta power, particularly in seizure disorder and status epilepticus (*p* < 0.1; Fig. 6**a,c,d(iii)**). This implies that patients with poorer overall health displayed beta power levels closer to normative trajectories.

Having characterized alterations in beta power across clinical groups, we next characterized effects of clinical groups of beta frequency. Figure 7 characterizes the alterations in beta frequency across our diagnostic groups, stratified by sex and brain lobes. Our analysis revealed that unadjusted beta frequency primarily showed beta slowing (Figure 7**a,c(i)**). This slowing was highly disorder and brain-lobe specific and was observed in seizure disorder, status epilepticus disorder, organic mental disorder with a low effect size (Cohen’s *d* < 0.2) (Fig. 7**a,c(ii)**). However, for dementia, unadjusted beta frequency showed beta acceleration with a medium effect size (Cohen’s *d* < 0.5). In contrast, adjusted beta frequency showed beta acceleration (Fig. 7**b,d(i)**). This effect was also highly disorder- and brain-lobe specific and was observed in organic mental disorder, dementia, and schizophrenia spectrum disorder with a low-to-medium effect size (Fig. 7**b,d(ii)**).

**Figure 7:**
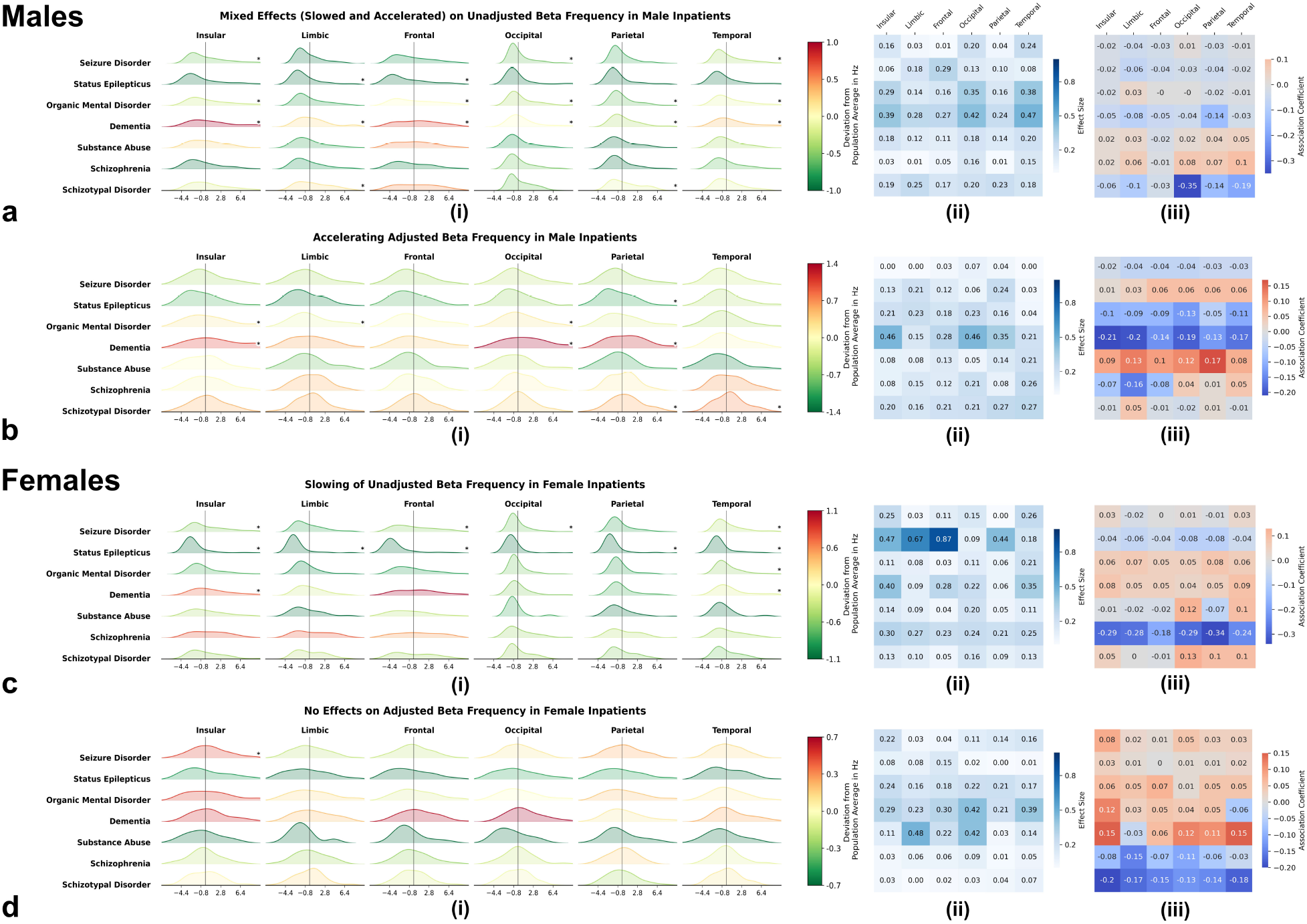
Alterations in Beta Frequency across Clinical Groups are Disorder and Brain-Region Specific. Panels a-d represents the characterization of beta frequency for clinical groups representing neurological disorders, stratified by sex, brain regions, and estimation method. **a-d(i)** Distributions of deviations in beta frequency from lifespan reference charts. Green distributions represent negative deviations indicating reduced beta frequency (beta slowing) while red distributions represent positive deviations indicating increased beta frequency (beta acceleration) in clinical groups, with statistical significance marked by (*). Unadjusted beta frequency primarily showed beta slowing (*p* < 0.1) across most conditions except dementia, where it accelerated. Adjusted beta frequency contrarily showed beta acceleration (*p* < 0.1). The effects for both unadjusted and adjusted beta frequency were highly disorder and brain-lobe specific. **a-d(ii)** Demonstrate the effect sizes of these deviations, with darker colors indicating stronger effects. The magnitude of these effects manifested with low-to-medium effect sizes, with status epilepticus and seizure dementia showing the highest effect sizes. **a-d(iii)** Display correlations between deviations in beta frequency and comorbidity scores with statistically significant correlations marked by circles. Negative correlations are shown as blue cells, while positive correlations are displayed in red cells. No statistically significant correlations were observed.

We also assessed the correlations between the deviations in beta frequency and comorbidity scores, but found no statistically significant correlations for both unadjusted and adjusted beta frequency (Fig. 7**a-d(iii)**). This analysis showed that unadjusted beta primarily showed slowing versus adjusted beta primarily showing acceleration in brain pathology, indicating the conflating effects of aperiodic 1/f activity in estimation of beta oscillatory parameters. Together, these findings demonstrate that beta oscillatory alterations in clinical populations are disorder-, region-, and method-specific.

## Discussion

In this study, we established the first comprehensive normative reference charts for beta rhythm parameters across the human lifespan and characterized neurological and neuropsychiatric disorders as deviations from these norms. A key methodological advance is the application of two complementary approaches: conventional band-pass filtering, which bridges inconsistent prior findings on lifespan beta changes, and spectral parameterization, which, for the first time in a normative lifespan context, isolates periodic beta oscillations from aperiodic 1/f activity. This dual-method approach directly quantifies the extent to which conventional beta measurements are confounded by aperiodic activity across development, a bias unaccounted for in existing normative frameworks. Our findings reveal that beta oscillatory parameters follow distinct, non-linear trajectories whose accurate characterization depends critically on separating aperiodic from periodic components. The divergence between conventional and aperiodic-adjusted measurements in both normative trajectories and disorder-specific deviations suggests that a substantial proportion of inconsistency in prior beta literature reflects aperiodic confounding rather than true oscillatory variability. The resulting age- and method-stratified framework enables more precise interpretation of beta parameters in research and clinical contexts.

### Beta Power shows a Tri-Phasic Lifespan Trajectory

Absolute and 1/f adjusted beta power, both showed a non-linear increase until mid-adulthood (age ~50 years), followed by a decrease in later years. On closer observation, however, this non-linear developmental trajectory demonstrated a tri-phasic pattern across the lifespan: non-linear increases throughout childhood and adolescence, continued increase in beta power reaching a peak around age 50, followed by decline through later life. The tri-phasic trajectory of beta power across the lifespan is consistent with the sequential maturation and eventual deterioration of GABAergic inhibitory circuits and myelinated thalamocortical networks. The sustained elevation into mid-adulthood distinguishes beta from alpha rhythms that stabilize after adolescence, suggesting distinct underlying maturational processes.

The initial increase in beta power during childhood and adolescence is consistent with the protracted maturation of GABAergic inhibitory circuits essential for generating beta oscillations. Beta rhythms emerge through the pyramidal-interneuron network gamma (PING) mechanism, where pyramidal neurons activate fast-spiking parvalbumin-positive (PV+) interneurons, which provide precisely-timed feedback inhibition creating rhythmic synchronization [49, 50]. PV+ interneurons undergo prolonged maturation in human cortex, with functional development continuing through adolescence into early adulthood [51]. Cortical GABA concentrations follow a parallel three-phase trajectory — developmental increases during childhood, stabilization in early adulthood, and decline with aging, that closely maps onto the observed beta power pattern [52]. Progressive myelination provides a third convergent process, with frontal white matter showing peak integrity around late 30s, optimizing the precise axonal conduction timing necessary for coherent oscillatory activity [53, 54].

The mid-adulthood peak around age 50 is consistent with optimal convergence of these maturational processes. From a functional perspective, middle age may represent a neurophysiological state characterized by enhanced network coordination and inhibitory control. Middle-aged adults show increased homogeneity in brain responses and cortical coupling compared to younger and older groups [55], and middle age is associated with increased strength of phase synchronization between beta and alpha oscillations relative to other life stages [56–58]. Resting beta may index network readiness and inhibitory control — processes highly engaged during the cognitively and physically demanding years of adulthood with compensatory mechanisms potentially boosting beta synchrony to offset subtle age-related inefficiencies before the decline seen in later life [18].

The later peak in females is consistent with two complementary processes. First, ovarian hormones sustain cortical GABAergic tone throughout the reproductive lifespan [59, 60], maintaining a higher GABAergic baseline in females than males from early adulthood onward [14, 61]. Estrogen is associated with regulation of GABAergic inhibitory tone through molecular mechanisms characterized in cortical and hippocampal circuits [62], and cortical GABA levels fluctuate measurably with ovarian hormone status in women [63]. Since beta oscillations are generated in part by GABAergic interneuron activity, this hormonally sustained inhibitory tone is consistent with the higher beta power observed in females across the lifespan [14, 61]. Second, the menopausal transition in the fifth decade is associated with a slowing of cortical GABA decline specifically in women [64, 65]; this decline is gradual rather than abrupt, and female beta power remains elevated for over a decade following menopause before falling, producing the observed ~15-year offset in peak age relative to males. Consistent with this pattern, sex differences in beta power amplify rather than converge with aging [66], reflecting progressive divergence of male and female GABAergic trajectories across the second half of the lifespan.

The subsequent late-life decline is consistent with convergent deterioration: reduced GABAergic interneuron function, progressive white matter degradation particularly in association cortices, and E/I balance shifts toward excitation dominance [32, 52]. Together, these findings indicate that beta power tracks the extended maturation and eventual deterioration of GABAergic inhibitory circuits across the lifespan, with mid-adulthood representing the peak of this maturational arc.

### Aperiodic Activity Confounds Beta Frequency Estimation

Unadjusted beta frequency showed relative stability during childhood, increases through adolescence, plateauing in mid-adulthood, then decline in later years. In contrast, aperiodic-adjusted beta frequency demonstrated a markedly different developmental pattern of decrease during childhood, a distinctive dip reaching minimum around ages 12-13 years, recovery through adolescence into the late 20s, and subsequent progressive decline beginning around age 30.

The divergence between unadjusted and adjusted beta frequency trajectories is a direct consequence of aperiodic spectral flattening across development artificially shifting apparent peak frequency. The aperiodic exponent progressively decreases with age throughout childhood and adolescence, representing systematic spectral flattening [32, 33]. This flattening is associated with artifactual increases in apparent peak frequency because relative power at higher frequencies is preserved as overall spectral slope decreases. When the aperiodic component is separated, the periodic component reveals network timing dynamics rather than spectral slope changes. The aperiodic component itself arises from temporal summation of excitatory and inhibitory postsynaptic currents with different decay kinetics, creating power-law spectral patterns that vary with E/I balance [28].

In contrast to the non-linear increase of beta power until mid-adulthood, adjusted beta frequency showed an early decline during childhood, a brief rebound in adolescence and early adulthood, and progressive slowing beginning around age 30, suggesting that the amplitude and frequency dimensions of beta oscillations follow partially distinct neurodevelopmental trajectories. The initial decline in adjusted beta frequency during childhood, occurring alongside rising beta power, may reflect early dominance of lower-frequency beta sub-bands before higher-frequency components emerge, consistent with evidence that high and low beta follow distinct developmental timelines in infancy [22] though direct evidence for this interpretation is lacking. The rebound in adjusted beta frequency through adolescence into early adulthood is consistent with fine-tuning of cortical circuits through synaptic pruning and myelination, which are associated with enhanced conduction velocity and synchrony during this period [67, 68]. This developmental pattern parallels the early rise in beta power, consistent with strengthening of inhibitory-pyramidal microcircuits that generate beta activity [9, 20, 69].

After early adulthood, power and frequency diverge. While beta power continues to increase into midlife, consistent with elevated GABAergic tone and pyramidal–interneuron synchronization [52], frequency begins to decline from around age 30, possibly reflecting sensitivity to subtle conduction delays and excitatory–inhibitory timing changes [70, 71]. Even as GABAergic tone remains sufficient to sustain strong oscillatory amplitude into midlife, subtle reductions in conduction velocity, dendritic integration, or excitatory–inhibitory timing may already be associated with slowing of beta cycle periodicity. Frequency therefore appears to capture an earlier signal of physiological slowing, whereas power may continue to reflect compensatory synchronization and sustained inhibitory efficacy into mid-adulthood.

From a functional standpoint, the combination of rising power and declining frequency during midlife may represent an optimal, but transient, balance: greater amplitude of oscillatory coordination but with slower cycle dynamics. This may support stable inhibitory control and network readiness during the cognitively demanding middle-adulthood years. Prior reports of increased phase synchronization between beta and alpha rhythms and greater homogeneity of cortical responses in midlife are consistent with this interpretation [55, 56, 72], suggesting that reduced frequency does not necessarily indicate dysfunction, but may rather reflect a reconfiguration of oscillatory regimes that maximizes efficiency during this stage.

n later years, both beta power and frequency decline, consistent with convergent age-related processes including interneuron dysfunction, white matter deterioration, and reduced GABA levels [13, 32, 58, 71]. Together, these findings indicate that beta power and frequency capture complementary aspects of inhibitory network maturation, efficiency.

### Beta Oscillations Show Disorder-Specific Alterations

Beta power showed disorder-specific alteration patterns, with most conditions including seizure disorder, status epilepticus, organic mental disorder, dementia, and substance abuse, showing reductions compared to normative trajectories, with effect sizes ranging from low to high depending on disorder category and brain region. Seizure disorder, status epilepticus, and organic mental disorders demonstrated the largest effects. This common pattern of beta power reduction across diverse neurological conditions [73] is consistent with shared vulnerability of GABAergic interneuron circuits and thalamocortical systems that generate beta oscillations [25, 50]. This reduction was present in both absolute and aperiodic-adjusted measures across most diagnostic groups, with the exception of schizophrenia spectrum disorders, which showed reduced absolute but increased aperiodic-adjusted beta power - a divergence undetectable using conventional band-pass approaches.

In epilepsy and seizure disorders, network hyperexcitability is associated with disruption of the E/I balance necessary for controlled oscillatory activity. GABAergic alterations observed in epileptic tissue, including interneuron loss or dysfunction, reduced GABA synthesis and availability, and altered receptor kinetics — are consistent with circuits shifted toward excitation dominance and reduced inhibitory precision required for beta synchronization [74, 75]. Thalamocortical circuit dysfunction further contributes, as beta generation depends on higher-order thalamic nuclei providing coordinated input to cortex [25]. In dementia and organic mental disorders, beta reduction is consistent with neurodegeneration affecting cortical microcircuits and modulatory systems. Cholinergic basal forebrain degeneration is associated with loss of acetylcholine modulation essential for thalamocortical oscillations, while PV+ interneurons show functional alterations including reduced activity and GABA release [76, 77].

Beta frequency alterations also showed disorder specificity. Unadjusted beta frequency primarily showed slowing in seizure disorder, status epilepticus, and organic mental disorder (Cohen’s *d* < 0.2), while dementia showed acceleration (Cohen’s *d* < 0.5). In contrast, aperiodic-adjusted beta frequency showed acceleration in organic mental disorder, dementia, and schizophrenia spectrum disorder (0.2 < Cohen’s *d* < 0.5). The opposing directions of unadjusted versus adjusted beta frequency changes across disorders offer a methodological explanation for previously inconsistent clinical reports: apparent beta slowing in conventional analyses may partly reflect aperiodic confounding rather than a true shift in oscillatory frequency, underscoring the importance of separating periodic from aperiodic components when interpreting beta frequency changes in clinical populations.

Schizophrenia spectrum disorders showed a pattern distinct from all other clinical groups: reduced absolute but increased aperiodic-adjusted beta power. This divergence likely reflects contributions from both disease pathophysiology and antipsychotic medication effects. PV+ interneuron dysfunction is a core feature of schizophrenia, manifesting as reduced inhibitory function rather than cell loss [78, 79]. This dysfunction is associated with paradoxical beta increases through several mechanisms: reduced inhibition is associated with enhanced pyramidal cell firing at certain frequencies, and altered thalamocortical connectivity with increased motor-thalamic coupling may amplify beta in motor circuits [80]. Antipsychotic medications are associated with increased beta band activity through basal ganglia-thalamocortical circuitry via mechanisms parallel to those observed in Parkinson’s disease [81]. The divergence between reduced absolute and increased adjusted beta power is consistent with confounding of periodic and aperiodic signals: schizophrenia is associated with altered E/I balance affecting aperiodic characteristics [82], while medications are associated with oscillatory changes through basal ganglia circuit effects [81]. Future studies comparing drug-naive and medicated patients are needed to disentangle disease and medication contributions to beta alterations in schizophrenia.

The inconsistencies that have hampered beta biomarker adoption in clinical practice stem from two compounding problems: beta changes in pathology substantially overlap with normal aging trajectories, and conventional band-power measures conflate periodic oscillatory changes with aperiodic 1/f shifts. Without age-appropriate normative baselines, the beta power reductions observed in seizure disorders, organic mental disorder, and dementia risk being conflated with normal age-related beta decline [11, 58]. Similarly, without aperiodic decomposition, the opposing frequency patterns observed here cannot be detected, leaving the mechanistic basis of beta frequency changes in pathology obscured [27, 83]. Our normative reference charts enable interpretation of individual patient EEG not only in terms of whether beta parameters deviate from age-appropriate expectation, but whether such deviations reflect periodic oscillatory changes or shifts in aperiodic 1/f activity, distinctions with potentially different mechanistic implications [84, 85]. By providing this distinction at the individual patient level, these reference charts offer a level of interpretive precision that conventional beta analyses cannot achieve.

### Strengths, Limitations and Future Directions

The primary strength of this study is establishing comprehensive lifespan reference charts for beta oscillatory parameters using both conventional and aperiodic-adjusted methods. The large sample size (N = 22,094) spanning ages 1-100 years provides robust normative trajectories accounting for non-linear developmental changes. The dual-method approach directly quantifies how aperiodic activity confounds traditional beta measurements across the lifespan, demonstrating this confound is substantial and varies with age. Clinical application demonstrates how deviations from age-appropriate norms reveal disorder-specific signatures that would be obscured without proper normative baselines.

Several limitations must be acknowledged. First, our reference population consists of outpatient EEGs, which may not perfectly represent healthy controls but provides a realistic benchmark for clinical translation, where comparison to idealized healthy controls is rarely feasible. Recent work has demonstrated the utility of normative modeling approaches that embrace population heterogeneity rather than requiring strict control criteria [85, 86]. Second, our cross-sectional design limits inference about individual developmental trajectories; the population-level patterns established here require longitudinal validation to confirm they reflect within-person change rather than cohort differences. Third, systematic medication information was unavailable for clinical groups, preventing analysis of pharmacological contributions. As the schizophrenia findings illustrate, medication effects can be substantial and may operate in opposing directions to disease effects. Future studies should systematically collect medication data, with comparison of drug-naive and medicated patients being particularly informative.

Key open questions raised by these findings include whether the adolescent dip in adjusted beta frequency represents a functionally significant developmental transition, whether longitudinal trajectories recapitulate the cross-sectional patterns established here, and whether aperiodic-adjusted beta parameters improve clinical classification accuracy over conventional band-power measures. More broadly, the importance of separating periodic from aperiodic activity is not specific to beta oscillations: future normative frameworks across all frequency bands should routinely provide both conventional and aperiodic-adjusted measurements to enable the kind of mechanistic disambiguation demonstrated here.

## Conclusion

This study established the first comprehensive lifespan normative reference charts for beta oscillatory parameters using a large-scale clinical EEG dataset spanning ages 1–100 years. Beta power followed a tri-phasic trajectory, increasing through childhood and adolescence, peaking around mid-adulthood, and declining through later life — a pattern consistent across sex, cortical lobes, and estimation methods. Beta frequency, in contrast, showed strikingly different developmental trajectories depending on whether aperiodic activity was accounted for, with unadjusted and adjusted measures tracing opposing patterns across the lifespan. A previously uncharacterized adolescent dip in adjusted beta frequency was identified as a genuine developmental feature, consistent across all brain lobes and both sexes. When applied to clinical populations, beta power reduction emerged as a common feature across seizure disorders, dementia, and organic mental disorders, while schizophrenia spectrum disorders showed a distinct pattern consistent with the interplay between disease pathophysiology and medication effects. These findings offer a partial reconciliation of contradictory prior reports, attributing a meaningful proportion of the inconsistency to two previously unaddressed factors: the inherently non-linear nature of beta developmental trajectories and systematic aperiodic confounding of conventional spectral measures. These reference charts provide a method-stratified normative framework for distinguishing age-related beta changes from pathology-driven deviations at the individual patient level.

## Data Availability

All data produced in the present study are available upon reasonable request to the authors. Only lobe averaged data will be made available.

## Data and Code Availability

Data supporting the findings of this study include de-identified participant-level, lobe-wise averaged values for several spectral parameters analyzed in the manuscript. During peer review, these data and the associated code for the analyses will be made available to editors and reviewers upon request. After publication, access to the minimum dataset will be provided upon reasonable request to the corresponding author, subject to applicable ethics approvals and data-sharing agreements required for human participant data.

